# Prognostic significance of compound physiology variables in oesophageal cancer

**DOI:** 10.1101/2020.04.16.20067769

**Authors:** Arfon G M T Powell, Catherine Eley, Alexandra H Coxon, Carven Chin, Damian M Bailey, Wyn G Lewis, South East Wales Oesophagogastric Cancer Collaborative

## Abstract

**Aims:** Objective identification of patient risk profile in Oesophageal Cancer (OC) surgery is critical. This study aimed to evaluate to what extent cardiorespiratory fitness and select metabolic factors predict clinical outcome.

**Methods:** Consecutive 186 patients were recruited (median age 69 yr. 160 male, 138 neoadjuvant therapy). All underwent pre-operative cardiopulmonary exercise testing to determine peak oxygen uptake 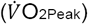, anaerobic threshold (AT), and ventilatory equivalent for carbon dioxide 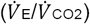. Cephalic venous blood was assayed for serum C-reactive protein (CRP), albumin, and full blood count. Primary outcome measures were Morbidity Severity Score (MSS), and Overall Survival (OS).

**Results:** MSS (Clavien-Dindo >2) developed in 33 (17.7%) and was related to elevated CRP (AUC 0.69, p=0.001) and lower *V*·O_2Peak_ (AUC 0.33, p=0.003). Dichotomisation of CRP (above 10mg/L) and *V*·O_2Peak_ (below 18.6mL/kg/min) yielded adjusted Odds Ratios (OR) for MSS CD>2, of 4.01 (p=0.002) and 3.74 (p=0.002) respectively. OC recurrence occurred in 36 (19.4%) and 69 (37.1%) patients died. On multivariable analysis; pTNM stage (Hazard Ratio (HR) 2.20, p=0.001), poor differentiation (HR 2.20, p=0.010), resection margin positivity (HR 2.33, p=0.021), and MSS (HR 4.56, p<0.001) were associated with OS.

**Conclusions:** CRP and *V*·O_2Peak_ are collective independent risk factors that can account for over half of OC survival variance.

## Introduction

Never before has there been such a variety of treatment modalities available, which in multimodal form can now cure as many as one-in-two oesophageal cancer (OC) patients.^1^ Oesophagectomy remains the primary therapeutic modality for radical and potentially curative treatment for patients with OC, but despite recent advances in anesthesiology and critical care it continues to carry significant inherent risk. Indeed, the 2018 UK National Oesophago- Gastric Cancer Audit ^2^ reported post-operative morbidity and mortality of 50% and 1.6% respectively. Current approaches to risk prediction comprise: clinical judgement, objective scoring systems such as the Portsmouth Physiological and Operative Severity Score for the enUmeration of Mortality and Morbidity (P-POSSUM)^3^, Oesophagogastric POSSUM (O-POSSUM)^4^, American Society of Anesthesiologists (ASA) physical status, Charleston Comorbidity Index, serum biomarkers, measures of cardiac function^5^, and the shuttle walk tests^6^. Their effectiveness in predicting surgical morbidity is relatively weak and measures to improve a clinician’s ability to predict outcome are needed. Cardiopulmonary exercise testing (CPET) is a non-invasive and dynamic procedure, which allows an individual’s cardiopulmonary fitness to be accurately measured. ^7^ CPET, in particular an anaerobic threshold <11 mL/kg/min, has been reported to predict post- operative morbidity and mortality in patients undergoing major abdominal surgery,^1,8–10^ yet, although well established in cardiothoracic surgery^11^, its application in the OC setting is limited.^10,12^

Cancer-related inflammation has been dubbed the seventh hallmark of cancer,^13^ and the systemic inflammatory response (SIR) is measured using cellular white cell counts (neutrophils, lymphocytes and platelets), humoral [C-reactive protein (CRP) and albumin] components. Derivative biomarkers (neutrophil-lymphocyte ratio (NLR), platelet-lymphocyte ratio (PLR), neutrophil-platelet score (NPS), and the modified Glasgow Prognostic Score (mGPS) have also been reported to be associated with survival.^14–16^ Despite emerging evidence that the SIR is associated with post-operative morbidity in colorectal cancer^17,18^, confirmatory evidence in OC is thin.

In light of the above, the present study examined to what extent select metrics of cardiorespiratory fitness and metabolic risk factors predict clinical outcome in OC patients scheduled for elective surgery. The hypothesis was that impaired cardiorespiratory fitness (CRF) and elevated CRP would predict patient morbidity and mortality. The primary outcome measures were post- operative morbidity severity, Overall Survival (OS), and Disease-Free survival (DFS)

## Methods

### Governance

Ethical approval was sought from the regional ethics committee, but a formal application was deemed unnecessary, because the study was considered to be a service evaluation of consecutively recruited patients, in whom consent had already been provided.

### Patients

#### Selection/staging

In order to test the hypotheses proposed in this study, a single cohort of patients diagnosed with oesophageal adenocarcinoma, between January 1, 2010 and August 31, 2018, was developed and included patients with radiological TNM stage I to III, deemed to have amenable to treatment with curative intent. All patients were managed by a multidisciplinary specialist team (MDT), with an interest in OC, and included clinical nurse specialists, gastroenterologists, surgeons, oncologists, radiologists, anaesthetists and pathologists.^19^ Management plans were individually tailored according to factors relating to both the patient and their disease. Patients were staged using computed tomography, endoscopic ultrasound, computed tomography positron emission tomography, and staging laparoscopy as appropriate. The South East Wales MDT treatment algorithms for OC have been described previously.^20,21^ The majority of these patients received 2 cycles of 80mg/m^2^ of Cisplatin and 1000mg/m^2^ of 5-FU for 4 days. A minority received 3 cycles of Epirubicin (50mg/m^2^), Cisplatin (60mg/m^2^) and 5-Fu (200 mg/m^2^) or Capecitabine (625mg/m^2^; ECF/X). Definitive chemoradiotherapy was offered to patients with localized squamous cell carcinoma and patients with adenocarcinoma deemed unsuitable for surgery because of disease extent and/or medical co-morbidity.^22,23^

#### Surgical intervention

The standard operative approach was subtotal Trans- Thoracic Oesophagectomy (TTO) as described by Lewis and Tanner.^24,25^ Trans-Hiatal Oesophagectomy (THO), as described by Orringer^26^, was used selectively in patients with adenocarcinoma of the lower third of the esophagus who had significant cardiorespiratory co-morbidity, clinical T1-3 N0 disease. A modified extended D2 lymphadenectomy (preserving pancreas and spleen where possible) was performed and the operative approach was open in 120 cases with 16 patients undergoing laparoscopic assisted surgery.

### Clinico-pathological Characteristics

Tumours were staged using the seventh edition of the AJCC/UICC-TNM staging system. Pathological factors were recorded from pathology reports issued at the time of surgery using AJCC/UICC-TNM staging system (seventh edition), and included tumour differentiation, number of lymph nodes with and without metastasis, and margin status.

Routine laboratory measurements of haemoglobin, whole white-cell count, neutrophil count, lymphocyte count, platelet counts, CRP, and Albumin on the day prior to surgery were recorded. Derivate measurements of systemic inflammation consisted of the NLR and PLR.^14^ The NLR and PLR were constructed by calculating the neutrophil to lymphocyte ratio and the platelet to lymphocyte ratio respectively.^14,27^

#### CPET testing

CPET followed American Thoracic Society/ American College of Chest Physicians recommendations.^11^ All patients performed a symptom limited CPET conducted on an electromagnetically braked cycle ergometer, and comprised 2 to 3min rest phase (to allow gas exchange variables to stabilise), 3 min unloaded cycling, then a ramped incremental protocol until volitional termination, and 2 to 5min recovery period. Ventilation and gas exchange was measured with a Medgraphics Ultima™ metabolic cart (Medical Graphics, St Paul, Minnesota, USA) with Breezesuite™ and Welch Allyn® (*Welch Allyn*, Inc., NY, USA) software as described previously.^12^

Heart rate, blood pressure, pulse oximetry, and 12-lead electrocardiogram were monitored throughout. The ramp gradient was set to 10 to 20 Watts based on the predicted *V*·O_2Peak_ from the age, weight, height, and sex of the patient in order to produce an exercise test of between 8-12 minutes duration ^28^. Prior to each test, the CPET equipment was calibrated against reference gases. The AT was determined using the V-slope method and confirmed by changes in ventilatory efficiency for oxygen 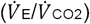. and end-tidal partial pressure values for oxygen (PET^O2^).^28^ The AT was validated independently by two experienced observers (IA and RD). *V*·O_2Peak_ was the highest V ^2^ achieved during the final 30 seconds of the test. The 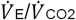 slope was measured at the AT. Test termination criteria included: request of patient, volitional fatigue, chest or leg pain, or electrocardiographic abnormalities determined by the consultant anesthetist. Multidisciplinary discussion and stratification of individual patient risk informed decisions regarding the planned post-operative level of care and invasive monitoring.

#### Morbidity/mortality

Operative morbidity was graded in accordance with the Clavien-Dindo Classification (CDC).^29–31^ Particular emphasis was placed on the incidence of morbidity of Clavien-Dindo grade >2, as this represented a complication requiring endoscopic, radiological or surgical intervention, in contrast with morbidity of lower grade requiring only pharmacological treatment.

#### Patient follow-up

Patients were followed up at 3-monthly intervals for the first year and 6 months thereafter. Investigations were undertaken sooner in the event that patients developed symptoms suggestive of recurrent disease. Surveillance was conducted for 5 years or until death, whichever was sooner.^32^ OS was calculated from time of diagnosis to the date of death. DFS was measured from the date of surgery until the date of recurrence or date of censoring. Causes of death were obtained from the Office for National Statistics via Cancer Network Information System Cymru (CaNISC). Recurrence patterns, which were characterised at the time of first diagnosis, were defined as loco- regional, distant (metastatic), or both. The date of recurrence was taken as the date of the confirmatory investigation.

## Statistical Analysis

Statistical analyses were performed using SPSS^®^ (IBM^®^ SPSS^®^ Statistics v25.0.0.0, IBM Corporation, Armonk, New York, USA) with extension R. Grouped data, that was not normally distributed based on Shapiro Wilks-W test, were expressed as median (interquartile-range) and non-parametric methods used. Receiver-operator-characteristic (ROC) analyses were employed to assess the predictive value of continuous variables with primary outcome measures and thresholds dichotomized for major morbidity as described by Youden et al.^33^For categorical variables, univariable and multivariable logistical regression analysis was used to identifying independent associations with major morbidity. Patient demographics were analyzed between the treatment modalities by means of Chi-Square χ^2^ or Mann Whitney *U* tests. Disease-free survival for all patients was calculated by measuring the interval from a landmark time of 6 months after diagnosis to the date of recurrence. This approach was adopted in previous randomized trials,^34^ to allow for the variable interval to surgery following diagnosis, depending on whether NeoAdjuvant Chemotherapy (NAC) was prescribed. As in these trials, events resulting in a failure to complete curative treatment, such as not proceeding to surgery, open and close laparotomy, palliative resection, in-hospital mortality and disease progression during NAC, were assumed to have occurred at this landmark time, to maintain the intention-to-treat analysis. Overall survival was measured from the date of diagnosis. Cumulative survival was calculated according to the method of Kaplan and Meier; differences between groups were analyzed with the log rank test. Univariable analyses examining factors influencing survival were examined initially by the life table method of Kaplan and Meier, and those with associations found to be significant (*p*<0·100) were retained in a Cox proportional hazards model using backward conditional methodology to assess the prognostic value of individual variables.

## Results

In total, 186 patients were identified who underwent potentially curative oesophagectomy for cancer. Twenty-five patients (13.4%) were deemed inoperable because of local tumour invasion. Of the patients undergoing surgical resection, 72 (44.7%) underwent a TTO, and 89 (55.3%) a THO. The median age for patients undergoing surgery was 69 years (IQR 64-73), 160 (86.0%) were male and 26 (14.0%) female. One hundred and thirty-eight (74.2%) patients underwent neoadjuvant chemotherapy. One hundred and one (54.3%) of patients developed a post-operative complication, with 33 (17.7%) being classified as major according to a Clavien-Dindo score of >2. There were five (2.7%) perioperative deaths. During follow-up, 36 patients (19.4%) developed cancer recurrence and 69 patients (37.1%) died. Median follow-up of survivors was 27 (range 7-60) months. One hundred and four (55.9%) patients were followed up for at least 5 years or death.

### Relationship between markers of the systemic inflammatory response, physiological variables, and MSS

The baseline and area-under-curve values for markers of the systemic inflammatory response and physiological variables can be found in table 1. There was no association between serum CRP and physiological parameters, with correlation values for anaerobic threshold (Spearman’s correlation coefficient (SCC) -0.080, p=0.286), *V*·O_2Peak_ (SCC -0.090, p=0.224), and 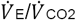 (SCC 0.093, p-0.210) were not statistically significant. Findings were similar for NLR and PLR (data not shown). Using a previously published dichotomization value of 10 mg/L^14^, 33 (17.7%) patients had a raised CRP.

**Table 1.** Association between markers of the systemic inflammatory response, physiological variables, and major morbidity

|  | Median (IQR) | Low / Normal / High (n)* | Area-Under-Curve<br>(95% Confidence interval) | p-value |
| --- | --- | --- | --- | --- |
| Serum variables |  |  |  |  |
| Haemoglobin | 133 (120-142) | 75 / 111 / 0 | 0.45 (0.34-0.55) | 0.318 |
| White Cell Count | 4.3 (5.2-7.8) | 11 / 173 / 2 | 0.60 (0.49-0.72) | 0.061 |
| Neutrophil Count | 3.9 (3.0-5.1) | 8 / 171 / 7 | 0.59 (0.48-0.70) | 0.104 |
| Lymphocyte Count | 1.6 (1.1-2.0) | 27 / 153 / 6 | 0.54 (0.44-0.65) | 0.426 |
| Platelet Count | 241 (203-291) | 5 / 175 / 6 | 0.51 (0.39-0.63) | 0.877 |
| **C-Reactive Protein** | **3.0 (1.0-7.0)** | **0 / 151 / 35** | **0.69 (0.60-0.79)** | **0.001** |
| Neutrophil-Lymphocyte Ratio | 2.64 (1.88-3.71) |  | 0.55 (0.44-0.65) | 0.415 |
| Platelet-Lymphocyte Ratio | 157 (124-223) |  | 0.46 (0.35-0.57) | 0.482 |
| CPEX variables |  |  |  |  |
| Anaerobic threshold | 11.5 (10.1-13.7) |  | 0.40 (0.30-0.51) | 0.082 |
| **$\dot{V}O_{2Peak}$** | **19.6 (16.4-23.5)** |  | **0.33 (0.23-0.43)** | **0.003** |
| $\dot{V}_E/\dot{V}_{CO_2}$ | 30.0 (27.0-33.3) | | 0.63 (0.52-0.73) | 0.024 |

\* Based on local thresholds

There was no difference between the median measurements of *V*·O_2Peak_, AT or 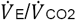, in patients with normal or high CRP respectively. The median value for CRP was 3 mg/L (interquartile range (IQR) 1-7). CRP was strongly associated with MSS (AUC 0.69 (95% CI 0.60-0.79), p=0.001, figure 1a). The median value for *V*·O_2Peak_was 19.6 mL/kg/min (IQR 16.4-23.5) and anaerobic threshold (AT) was 11.5 mL/kg/min (IQR 10.1-13.7) (table 1). Using the Youden index, the optimum dichotomization threshold for *V*·O_2Peak_ was 18.6 mL/kg/min (figure 1b), and AT was 11.5 mL/kg/min with 43.5% and 48.9% of patients considered to have low measurements respectively. This gave sensitivity and specificity of 69.7% and 62.1% respectively for *VO*^*2Peak*,^ and 69.7% and 53.4% respectively for AT. Total morbidity (CD>1) and operative mortality rates were 53.1% and 1.2% for low *V*·O_2Peak_ and 60.2% and 2.3% for low AT respectively.

**Figure 1.**
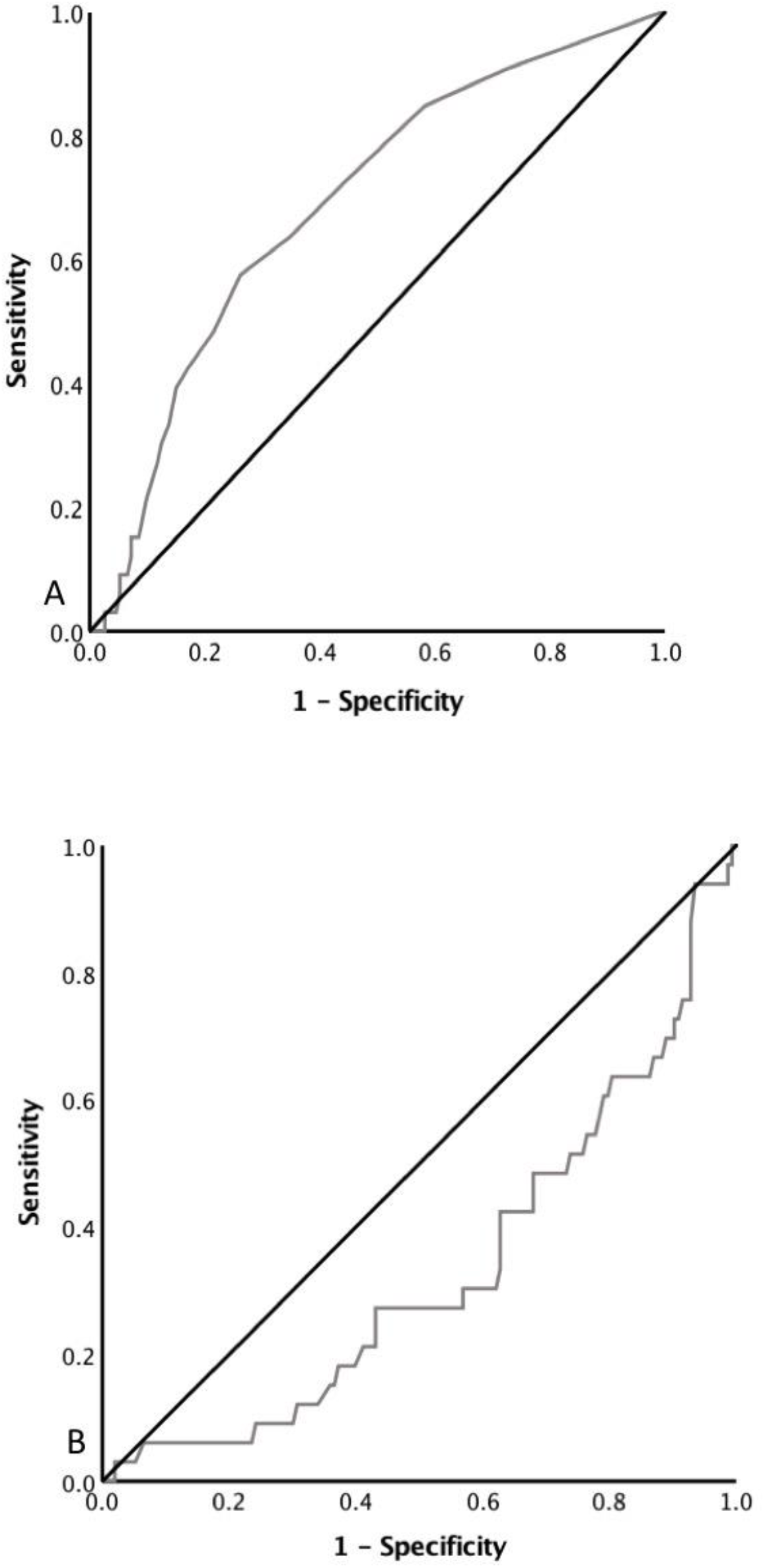
Predictive value of (A) CRP, (B) *V*·O_2Peak_, and major morbidity.

To adjust for potential confounders, a binary logistical regression model was developed to include the clinical factors available to the MDT at the point of deciding on definitive treatment (table 2). On univariable binary logistical regression analysis, only CRP (p=0.022), *V*·O_2Peak_ (p=0.001), and AT (p=0.069), were associated with major morbidity. On multivariable logistical regression analysis, CRP (odds ratio (OR) 4.01 (95% CI 1.66-9.66), p=0.002) and *V*·O_2Peak_ (OR 3.74 (95% CI 1.62-8.65), p=0.002) were independently associated with major morbidity. A composite score was developed to determine if major morbidity could be predicted with greater accuracy. The Combined Inflammatory and Physiology Score **(CIPS)** ranged from zero to two. Patients with a normal CRP and *V*·O_2Peak_ were given a score of zero (low), a score of one (intermediate) was given to patient if either the CRP was high or *V*·O_2Peak_ was low, and a score of two (high) was given to patients with both an elevated CRP and low *V*·O_2Peak_. This resulted in 88 (47.3%), 80 (43.0%), and 18 (9.7%) patients being classified with CIPS of low, intermediate, and high respectively. The major morbidity rate was 9.1% (n=8), 17.5% (n=14), and 61.1% (n=11) in the low, intermediate, and high **CIPS** cohorts respectively (p<0.001). A stepwise association between advancing **CIPS** and major morbidity was observed. Compared with the low CIPS cohort, OR was 2.12 (95% CI 0.84-5.36) for intermediate, and 15.71 (4.76-51.87, p<0.001) for high CIPS.

**Table 2.** Univariable and multivariable analysis of pre-operative factors associated with major morbidity

|  | **Univariable** | **p-value** | **Multivariable** | **p-value** |
| --- | --- | --- | --- | --- |
|  | **Odds ratio (95% CI)** |  | **Odds ratio (95% CI)** |  |
| Age (Years) (<65 / 66-75 / >75) | 1.29 (0.73-2.26) | 0.386 |  |  |
| Gender (Female / Male) | 1.29 (0.42-4.02) | 0.658 |  |  |
| Differentiation (Well-moderate / Poor) | 0.56 (0.26-1.22) | 0.144 |  |  |
| cTNM (1 / 2 / 3 / 4) | 0.844 (0.46-1.56) | 0.590 |  |  |
| Neoadjuvant therapy (No / Yes) | 0.62 (0.28 – 1.37) | 0.239 |  |  |
| Surgical approach (THO / TTO) | 1.11 (0.51-2.40) | 0.792 |  |  |
| C-reactive Protein (Normal / High) | 2.85 (1.16 – 6.98) | 0.022 | 4.01 (1.66-9.66) | 0.002 |
| V'O<sub>2Peak</sub> (<18.6 / ≥18.6) | 3.92 (1.76 – 8.73) | 0.001 | 3.74 (1.62-8.65) | 0.002 |
| Anaerobic Threshold (<11.5 // ≥11.5) | 2.06 (0.95-4.50) | 0.069 |  | 0.735 |

### Relationship between clinicopathological factors and OS

The relationship between clinicopathological factors and OS can be found in table 3. The cumulative OS for **CIPS** and MMS can be found in figure 2.

**Table 3.** Univariable and multivariable analysis of factors associated with OS

|  | Univariable |  | Multivariable |  |
| --- | --- | --- | --- | --- |
|  | HR (95% CI) | p-value | HR (95% CI) | p-value |
| Age (<65 / 66-75 / >75 yr) | 1.11 (0.78-1.58)) | 0.565 |  |  |
| Gender (Female / Male) | 1.11 (0.57-2.18) | 0.753 |  |  |
| Operative Approach (TTO / THO) | 0.60 (0.34-1.04) | 0.069 |  | 0.622 |
| CRP (Normal / High) | 1.92 (1.12-3.30) | 0.017 |  | 0.513 |
| V'O<sub>2Peak</sub> (Normal / Low) | 1.54 (0.95-2.48) | 0.079 |  | 0.720 |
| Neoadjuvant therapy (No / Yes) | 1.00 (0.98-1.02) | 0.936 |  |  |
| Pathological TNM stage (0 / 1 / 2 / 3 / 4) | 2.99 (1.99-4.48) | <0.001 | 2.20 (1.37-3.55) | 0.001 |
| Differentiation (Well-moderate / Poor) | 2.92 (1.75-4.88) | <0.001 | 2.20 (1.21-4.00) | 0.010 |
| CRM Margin (Negative / Positive) | 1.75 (1.44-2.13) | <0.001 | 2.33 (1.14-4.77) | 0.021 |
| Lymph Node Yield (<15 / ≥ 15) | 1.62 (0.93–2.81) | 0.088 |  | 0.746 |
| Major Morbidity (No / Yes) | 2.09 (1.22-3.59) | 0.007 | 4.56 (2.35-8.84) | <0.001 |
| Combined Inflammatory and Physiology Score (0 / 1 / 2) | 1.68 (1.17-2.42) | 0.005 |  | 0.934 |

**Figure 2.**
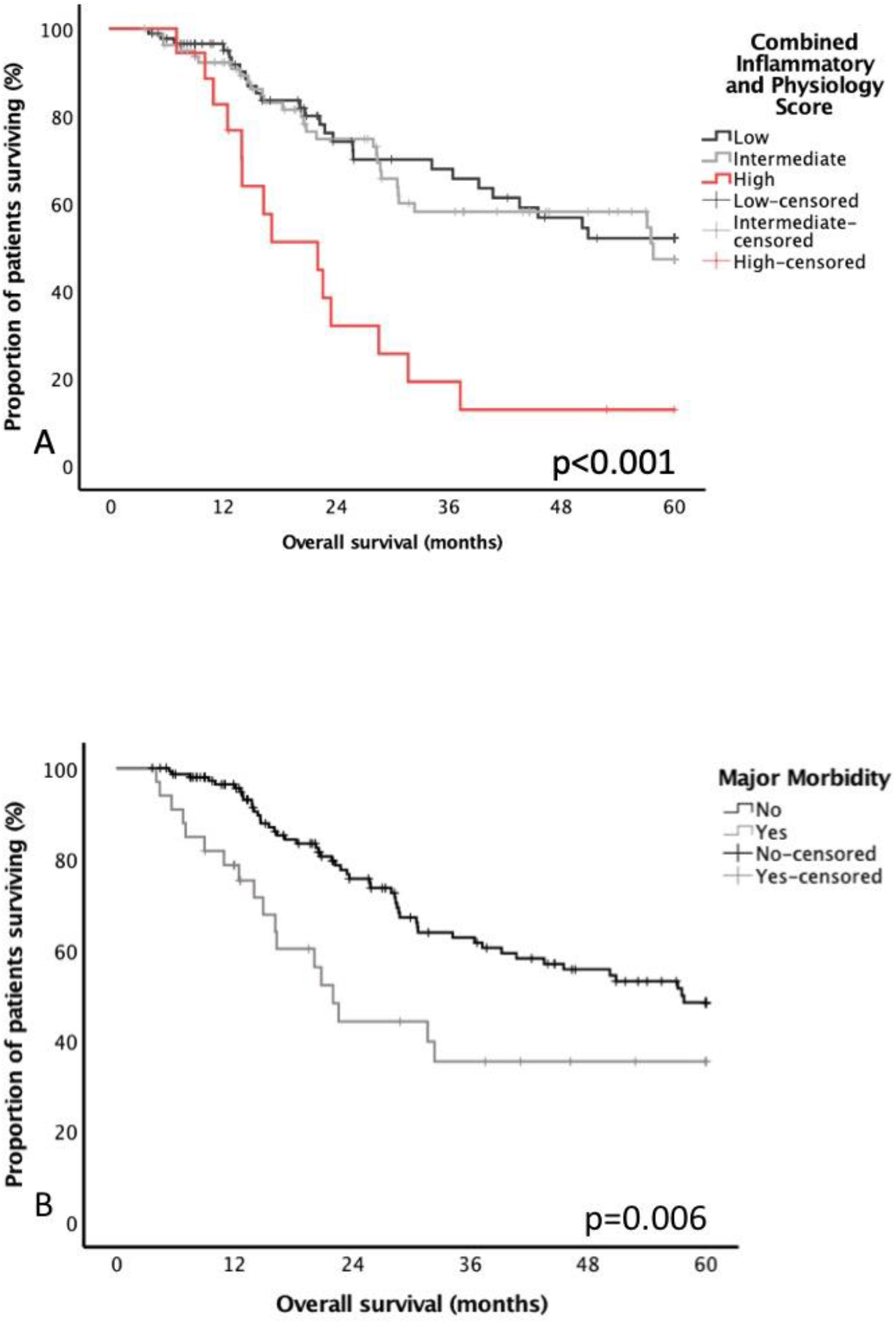
Cumulative OS related to (A) Combined Physiological and Inflammatory Score and (B) Major Morbidity.

### Relationship between clinicopathological factors and DFS

The relationship between clinicopathological factors and DFS can be found in table 4.

**Table 4.** Factors associated with Disease-Free Survival

|  | Univariable |  | Multivariable |  |
| --- | --- | --- | --- | --- |
|  | HR (95% CI) | p-value | HR (95% CI) | p-value |
| Age (<65 / 66-75 / >75 yr) | 1.09 (0.68-1.76) | 0.722 |  |  |
| Gender (Female / Male) | 2.06 (0.63-6.73) | 0.230 |  |  |
| Operative Approach (TTO / THO) | 1.17 (0.58-2.35) | 0.657 |  |  |
| CRP (Normal / High) | 1.03 (0.40-2.65) | 0.949 |  |  |
| V'O<sub>2Peak</sub> (Normal / Low) | 1.21 (0.63-2.32) | 0.571 |  |  |
| Neoadjuvant therapy (No / Yes) | 0.98 (0.91-1.05) | 0.544 |  |  |
| Pathological TNM stage (0 / 1 / 2 / 3 / 4) | 2.33 (1.51-3.57) | <0.001 | 2.08 (1.24-3.50) | 0.005 |
| Differentiation (Well-moderate / Poor) | 3.21 (1.60-6.45) | 0.001 |  | 0.055 |
| CRM Margin (Negative / Positive) | 2.06 (1.07-3.98) | 0.031 |  | 0.849 |
| Lymph Node Yield (<15 / ≥ 15) | 1.31 (0.67–2.55) | 0.428 |  |  |
| Major Morbidity (No / Yes) | 1.57 (0.71-3.45) | 0.262 |  |  |
| Combined Inflammatory and Physiology Score (0 / 1 / 2) | 1.16 (0.67-2.01) | 0.603 |  |  |

## Discussion

The principal finding of this study was that metabolic measures of the Systemic Inflammatory Response (SIR) together with physiological measures of cardiorespiratory fitness (*V*·O_2Peak_), were independently associated with major complications after potentially curative oesophagectomy for cancer, supporting the primary hypothesis. Major operative morbidity was three-fold higher in patients with high CRP and poor CRF, with a sensitivity and specificity of 39.4% and 85.6% for CRP and 69.7% and 62.1% for *V*·O_2Peak_, respectively. Moreover, combining these parameters established a novel composite risk score **(CIPS)**. Based on a **CIPS** of two, no fewer than 11 of 18 patients (61.1%) developed major morbidity, compared with eight (9.1%) with a **CIPS** of zero. Similarly, patients with a **CIPS** of zero experienced five-year OS that was more than two-fold greater at 50%, compared with 18% in patients with a **CIPS** of two.

Previous reports have contended that the SIR is closely associated with post-operative complications in colorectal cancer^35^. Richards *et al*, reported that CDC morbidity rates were 28% and 44% in patients with a modified Glasgow Prognostic Score of zero and two respectively. The pathophysiological cause for this association is unclear but likely relates to the underlying aetiology of the SIR, with aggressive tumour biology and individual patient CRF likely contributing factors. In the presence of cardiovascular disease, diabetes, poor diet, obesity, and smoking have all been reported to be associated with elevated CRP and poorer prognosis^36^. Moreover, modification of these lifestyle factors resulted in SIR resolution. Nevertheless, the data here did not show any correlation between raised CRP and physiological factors, arguably supporting the concept that an activated SIR prior to surgery has a multifactorial aetiology. We have previously demonstrated that a low *V*·O_2Peak_ is independently associated with major morbidity following oesophagectomy for OC, and measures to attenuate the SIR and poor CRF have the potential to reduce morbidity and prolong survival^37^. Unfortunately, data on lifestyle factors and anti-inflammatory use were not available for analysis and their associations with pre-operative inflammatory and physiological factors is worthy of further study.

These findings raise the possibility of whether a focused programme of prehabilitation combined with measures to attenuate the SIR, may reduce peri-operative complications, and enhance survival. Barberan-Garcia *et al* reported a randomised control trial (RCT) of prehabilitation in elective major abdominal surgery^38^ and showed that prehabilitation reduced postoperative complications by 51%. Unfortunately, approximately 60% of patients undergoing oesophagectomy will develop post-operative morbidity, most related to compromised respiratory function^39^. Minnella *et al*, of Montreal, Canada, reported a randomised control trial of respiratory function prior to and following surgery^40^. Prehabilitation was associated with higher functional capacity before surgery (mean [SD] 6MWD change, 36.9 [51.4] vs. −22.8 [52.5] m; p < .001), which was maintained into the post-operative period (15.4 [65.6] vs. −81.8 [87.0] m; p < 0.001). Results, which are very promising for a patient cohort whose functional and oncological outcomes, are relatively poor.

Based on the prevailing evidence, it appears that prehabilitation programmes including measures to attenuate the SIR are desirable if not urgently needed. What remains to be clarified is what optimum method of SIR attenuation is most suited to cancer patients, carrying significant pre-existing morbidity, and facing complex major surgery. Moreover, will patients with a CIPS>0 derive the most benefit from these attenuation measures? Although it would seem reasonable to incorporate anti-inflammatory medication into a prehabilitation care package, emerging evidence suggests that this is not without risk. A meta-analysis of Non-steroidal anti-inflammatory use in colorectal surgery suggested an increased risk of anastomotic leak (OR 1.96)^41^. A similar finding was also observed in patients undergoing oesophago-gastrectomy (OR 5.24)^42^. Yet these findings remain controversial, indeed McSorley et al reported that two doses of peri-operative dexamethasone, reduced the post-operative inflammatory response and complication rate in patients undergoing colectomy for cancer^43^. Therefore, it may be that patients with a CIPS>0, which accounted for 75% 0f all major morbidity in this study, will derive the most benefit from SIR attenutation. The main causes of major post-operative morbidity in patients undergoing oesophagectomy are related to sepsis, namely respiratory failure and anastomotic leak. Given that wound healing relies heavily on the inflammatory response, it may be prudent to omit NSAIDs and other anti-inflammatory medication from the prehabilitation bundle. The findings by Sattar and colleagues that lifestyle modification reversed the SIR also support this concept ^36^. It is possible that a proportion of patients with a SIR may not respond to prehabilitation, and therefore constitute a self-selecting cohort that may benefit from pharmacotherapy. Adequately powered studies to examine the effect of prehabilitation on SIR modulation are clearly needed to guide prehabilitation programme development.

Validating these results in an appropriately powered independent cohort would help integrate this novel combined inflammation-physiological score into a modified OG cancer treatment pathway, but a number of potential inherent and hypothetical limitations, related to the methodology of the present study mean that the findings must be interpreted with caution. The patient cohort constituted a highly selected group (most had undergone a potentially curative R0 oesophagogastrectomy), and were not universally representative of patients diagnosed with oesophageal cancer.^44^ Moreover, clinical access to CPET remains limited, with the most recent literature reporting that only 32% of UK hospitals have ready access to this applied multidisciplinary physiological asset^45^. Data relating to blood loss and operation time was not collected for this study and therefore is not available for analysis. Nevertheless, it is unlikely that these are considerable confounders for SIR and physiology variables in predicting post-operative morbidity. CPET assessment was first introduced in 2010 and therefore the follow-up period is slightly immature, nevertheless, strong statistical signals are identified and CIPS is worthy of validation in an independent cohort. In contrast, the study has several strengths, benefiting from robust follow-up data - two thirds of patients followed up for at least 5 years or death - with accurate causes and dates of death obtained from the office of national statistics. A NHS laboratory using standardized techniques performed the serum measurements and reporting of pathology specimens, and therefore re-examination of these findings in another comparable OC patient cohort, should be eminently pragmatic. Moreover, the patients were recruited from a consecutive series of patients diagnosed with OC, from a single UK geographical region, all treated by the same specialist MDT, using a standardized staging algorithm and team-based operative techniques, with international audited and published quality control.^19^

In conclusion, CRP and *V*·O_2Peak_ are important in the risk assessment of patients undergoing oesophagectomy for cancer. Combining these variables into a novel prognostic score improved the predictive accuracy further. Refining cardiopulmonary fitness by using a multimodal prehabilitation treatment bundle may also attenuate the SIR, potentially reducing post- operative morbidity, improving quality of life, and long-term survival, without the need for anti-inflammatory medication.

## Data Availability

Queries and requests for data can be made to the corresponding author

## Notes

### Competing Interest Statement

The authors have declared no competing interest.

### Funding Statement

None

